# Incidence and Predictors of Neuropsychiatric manifestations following a Traumatic Brain Injury in Referral Hospitals in Dodoma, Tanzania: a protocol of a prospective longitudinal observational study

**DOI:** 10.1101/2023.08.20.23294327

**Authors:** Suluma Aslan, Azan Nyundo

**Author notes:** **Corresponding author** Azan Nyundo P O Box 395 Dodoma 1 TIBA STREET, 41218 IYUMBU, DODOMA TANZANIA.

## Abstract

**Introduction:** Traumatic Brain Injury (TBI) is any injury to the brain resulting from an external force leading to complications. TBI affects 27-69 million yearly, with high incidence in Africa and LMICs, and is attributed to motor traffic accidents. Within three to six months following moderate-to-severe TBI, the relative risk of any psychiatric disorder is significantly higher than in the general population. Post-TBI neuropsychiatric disorders include; depression with a prevalence of over 50%, apathy up to 72%, Post-traumatic stress disorder (26%), anxiety (9%), manic symptoms (5 - 9%) and psychosis (3 to 8%).

This study aims to determine the incidence and predictors of post-TBI neuropsychiatric manifestations among patients admitted at Referral hospitals in Dodoma.

**Methods and analysis:** This is a prospective longitudinal observational study in which patients admitted after moderate to severe TBI will be recruited after obtaining informed consent. Patients will be followed for six months; the diagnostic MINI International Neuropsychiatric Interview (M.I.N.I) tool will assess psychiatric disorders, and severity and progression of symptoms will be assessed using PHQ-9 for depressive symptoms, GAD7 for anxiety symptoms, PCL-5 for Post-traumatic Stress Disorders (PTSD), MoCA for cognitive assessment, AES for apathy and YMRS for manic symptoms at one, three and six months. The analysis will use logistic regression to determine the association between predictors and neuropsychiatric outcomes.

**Ethics and dissemination:** The ethical clearance has been secured from the institutional Research Review committee and ethical committee of the University of Dodoma with the reference number MA.84/261/12. The respective authorities provided permission to conduct the study within the premises of BMH and DRRH.

**Strength and limitation:** The diagnostic tool “International Neuropsychiatric Interview (M.I.N.I),” is highly reliable and sensitive as it utilizes both DSM-5 and ICD-10 criteria, and screening tools there are used have good validity and reliability in assessing the severity progression of neuropsychiatric symptoms.

The longitudinal prospective nature of the study offers a strong temporal association of the incidence, progression and associated factors of neuropsychiatric manifestations post-TBI.

The study excluded patients with a known history of psychiatric diagnosis that could influence the recurrence relapse of the present illness and the existence of another new psychiatric diagnosis after TBI.

## INTRODUCTION

Of 7.3 billion people, 27 to 69 million are expected to suffer from TBI annually(Dewan et al., 2019; James et al., 2019). The majority of TBI cases occur in low- and middle-income countries (LMIC), comprising 85% of the world’s population and roughly 90% of all injury-related fatalities(Rao, Koliatsos, Ahmed, Lyketsos, & Kortte, 2015).

TBI is predicted to significantly burden Africa, with around 6 and 14 million additional cases by 2050(Wong, Linn, Shinohara, & Mateen, 2016). A study at Muhimbili Orthopedic Institute (MOI) in Dar es Salaam, Tanzania, reported that TBI is most commonly caused by traffic accidents (Boniface, Lugazia, Ntungi, & Kiloloma, 2017).

The severity of TBI is determined by the Glasgow Coma Scale (GCS), with a maximum score of 15 scores range categorized as mild (13-15), (9-12) as moderate and (≤ 8) as severe, the scale combines eye, motor, and visual responses to rate levels of consciousness)(*GCS.Pdf*, n.d.; Grubaugh, 2014).

TBI can result in several neuropsychiatric (NPS) disorders, including; depression, with an estimated prevalence of over 50% in one year, which is the primary cause of disability among those who have experienced a traumatic brain injury (TBI)(Moldover, Goldberg, & Prout, 2004). Furthermore, prior history of depression and alcohol dependence are risk factors for post-TBI depression(Bombardier et al., 2010).

With a 20 to 72 per cent prevalence (Al-Adawi et al., 2004), apathy is among the most common neuropsychiatric outcomes following traumatic brain injury(Starkstein & Pahissa, 2014). Based on Glasgow Outcome Scale, severe disabilities after TBI are likely to exhibit apathetic behaviour (Van Reekum, Stuss, & Ostrander, 2005). Closely mimicking apathy are the cognitive deficits that may be caused by primary brain injury or secondary impairments, including sleep disturbances(Ahmed et al., 2017). Potential causes of cognitive impairment post-TBI include pre-TBI cognitive impairment, TBI severity, involvement of specific brain regions, the damage mechanism, genetics, and advanced age (Cristofori & Levin, 2015).

Other neuropsychiatric manifestations of TBI include anxiety disorders, psychosis, manic symptoms and neurocognitive disorders(Ahmed et al., 2017). The rate of anxiety disorder ranges between 19 and 50 per cent are reported at distinct post-injury durations(Alway, Gould, Johnston, McKenzie, & Ponsford, 2016; Balan, Walz, Diaz, & Schwarzbold, 2021). During six months period following TBI, 26.8% screen positive for PTSD, which is associated with functional impairment, post-concussive symptoms, cognitive decline related to poor performance in visual processing and mental flexibility and overall decline in life satisfaction(Haarbauer-Krupa et al., 2017). While PTSD is more likely to result from psychologically upsetting events, such as memory of the traumatic events and early post-traumatic stress symptoms(Stein et al., 2019), neuroanatomical factors, including Hippocampus volume loss, amygdala hyperactivity, and hypo activity in the ventromedial prefrontal cortex(vmPFC) are all associated with PTSD after TBI (Motzkin, Philippi, Wolf, Baskaya, & Koenigs, 2015).

Manic symptoms are also reported after TBI, with a prevalence of 5-9% (Koponen, Taiminen, Hiekkanen, & Tenovuo, 2011), with focal brain injuries in the frontal lobe being implicated in the genesis of post-TBI manic symptoms (Satzer & Bond, 2016). Compared to the general population, newly diagnosed Bipolar I Disorder are 1.5 times more likely to have experienced a head injury in the past five years(Mortensen, Mors, Frydenberg, & Ewald, 2003).

As for psychotic symptoms, 3-8% of TBI patients get psychotic illnesses after their injury (Vaishnavi, Rao, & Fann, 2009). Risk factors for post-traumatic psychosis include male gender, pre-injury neuropsychiatric problems, family history of schizophrenia, severity of the brain damage, involvement of the temporal and frontal lobes, left hemisphere laterality, EEG abnormalities, post-traumatic epilepsy, and pre-TBI cognitive impairment(Schwarzbold et al., 2008).

Although the incidence and predictors of neuropsychiatric manifestations of TBI are widely reported elsewhere, the knowledge gap is wider in sub-Saharan settings. To the best of our knowledge, there are only a few published studies in Africa regarding post-TBI neuropsychiatric manifestations(Bangirana et al., 2019; Joosub, Cassimjee, & Cramer, 2017), but none has been published in Tanzania. Given the severity of neuropsychiatric disorders and related short and long-term complications, early diagnosis and prompt intervention can prevent long-term disability from TBI (Jesse R. Fann, MD, 2004; Kohnen, Lavrijsen, Akkermans, Gerritsen, & Koopmans, 2020), through specific tailor-made recovery and rehabilitation programs suited to the patient’s needs(Bangirana et al., 2019).

The study aims to determine cumulative incidence and factors associated with neuropsychiatric manifestations after TBI among patients admitted at referral Hospitals in Dodoma region, Tanzania.

### Specific aims

1. To describe baseline neuropsychiatric disorders at one month following TBI in referral hospitals in Dodoma, Tanzania.
2. To determine predictors of baseline neuropsychiatric manifestation in TBI patients in referral hospitals in Dodoma.
3. To determine the cumulative incidence of neuropsychiatric disorders at three and six months following TBI in referral hospitals in Dodoma, Tanzania.
4. To determine factors associated with any neuropsychiatric disorder at three and six months following TBI in referral hospitals in Dodoma, Tanzania.

## MATERIALS AND METHODS

### Study design

This is a prospective longitudinal observational study.

### Study area

Study will be conducted at the referral hospitals in Dodoma; The Benjamin Mkapa Hospital and Dodoma Regional Referral Hospital. Dodoma is the capital city of Tanzania located in the central part of the country, serving approximately three million people from Dodoma and nearby regions (Statistics, 2022). The bed capacities of the two hospitals are 400 and 480 for DRRH and BMH, respectively. Based on local data, these hospitals receive a total number of 7 to 10 patients of moderate to severe TBI per week. The hospitals have surgeons and neurosurgeon qualified for procedures needed after TBI. In case of neuroimaging, CT scan and MRI are performed at BMH. Patients will be found in both hospitals’ general surgery ward in DRRH and neurosurgery ward in BMH and ICU.

### Study population

This study involves all patients aged 18 years and above admitted at DRRH or BMH after moderate or severe TBI within seven days after TBI. Some of these patients are brought directly to these hospitals, and others are referred from district hospitals surrounding Dodoma region.

### Study duration

This study will take 12 months from June 2023 to August 2024, in which the enrollment will be done for four to six months, and every patient will be followed for 6 months.

### Inclusion criteria

1. Patients with age between 18 years and above admitted after their first traumatic brain injury
2. Patients who scored less than 13 in GCS
3. Patients with the capacity to provide informed consent, or close family member or custodian for those who cannot.

### Exclusion criteria

1. Patients with a known CNS diagnosis such as CNS tumours, or any space occupying lesion of the brain.
2. Patient with recent history of severe stroke.
3. Patient with past history of psychotic disorders like Schizophrenia spectrum disorder prior to TBI.

### Sample size

We use Krejcie and Morgan formula to calculate the sample as follows;

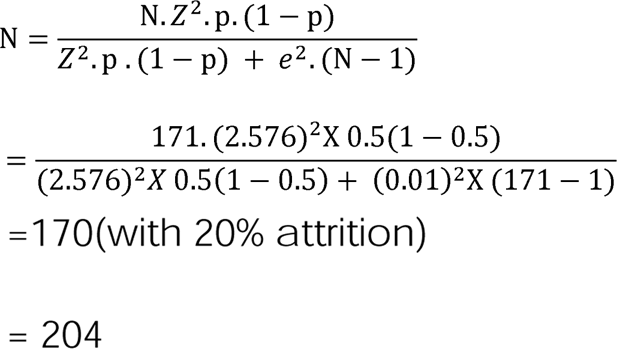

Where n - Calculated sample; N – Population =171 from (Bangirana et al., 2019) Z - Normal standardized variable associated with the confidence level=2.236; p-True probability of the event=50%(because there was no prev. literature) e - sample error=0.01

#### Sampling procedure

A consecutive sampling method will be used to recruit all the patients who meet inclusion criteria found in the surgical ward at DRRH and BMH surgical ward and ICU. This will be done by taking all the available patients who fit the inclusion criteria until the desired sample is reached.

### Data collection

#### At baseline

A Principal investigator and research assistant will handle the consent procedure. Once the study details are explained and understood, the patient or the custodian will be asked to sign the informed consent with a signature or thumbprint. For follow-up, at least two contacts will be recorded, including participants’ contact and caregiver or close family or friend contacts. Patient’s information will be entered into Open Data Kit (ODK) using an android mobile device where social demographic characteristics, including age, sex, education level, and occupation, will be collected. A history of substance use and history of being intoxicated with alcohol prior to TBI will also be documented.

TBI clinical presentation will be documented, including the severity of the TBI measured by using the Glasgow Coma Scale (GCS), which ranges from mild (14-15), moderate (9-13), and severe (8-3) immediately following an injury. Associated clinical presentations including seizures, speech difficulties and confusion and laterality will be recorded. Nature of the trauma will be classified open or close, penetrative or non-penetrative. Cause of injury will be recorded as Road Traffic Accident (RTA), (motor cycle, motor vehicle), falls, and hit by objects or sports. Type of TBI lesions including hematoma, contusion, intra cerebral hemorrhage, subarachnoid hemorrhage, diffuse axonal injury will be documented after being confirmed by CT scan or MRI as part of clinical workup. These patients will be followed up and assessed for neuropsychiatric manifestations at one, three and months.

#### At one month

Baseline assessment of the neuropsychiatric manifestations, including depression, anxiety, bipolar, apathy, psychosis and neurocognitive symptoms, will be assessed using standardized diagnostic and screening tools(See the data collection instrument).

#### At three and six months

The cumulative incidence of neuropsychiatric disorders will be assessed. The onset of a new neuropsychiatric disorder will be identified using the screening tools used at baseline. The tools will identify significant change in severity of neuropsychiatric symptoms, defined as either a significant reduction in symptoms that the criteria is no longer met or an increase in symptoms severity that a new diagnosis is currently met.

## OUTCOME MEASUREMENTS

### Definition and measurement of variables

**Aim 1 study variables;** these variables address baseline neuropsychiatric disorders at one month following TBI. The presence of neuropsychiatric manifestations will be measured using screening and diagnostic tools (See table 1, in the appendix for a listing and brief description of the variables for aim 1).

**Table 1.**
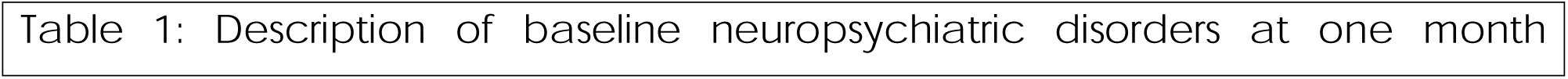
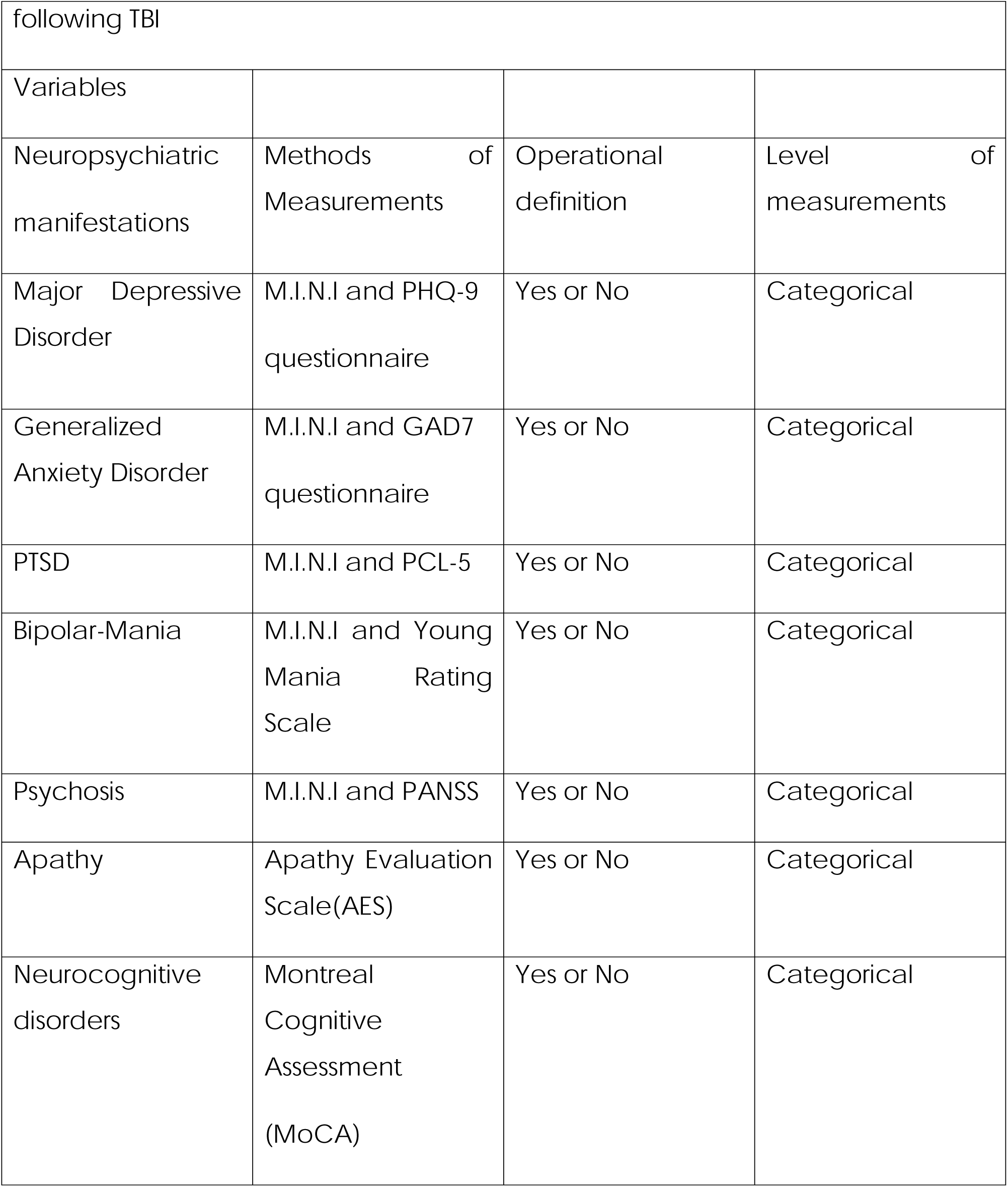
Aim 1.

|  |
| --- |
| Table 1: Description of baseline neuropsychiatric disorders at one month |

|  |  |  |  |
| --- | --- | --- | --- |
| following TBI |  |  |  |
| Variables |  |  |  |
| Neuropsychiatric manifestations | Methods of Measurements | Operational definition | Level of measurements |
| Major Depressive Disorder | M.I.N.I and PHQ-9 questionnaire | Yes or No | Categorical |
| Generalized Anxiety Disorder | M.I.N.I and GAD7 questionnaire | Yes or No | Categorical |
| PTSD | M.I.N.I and PCL-5 | Yes or No | Categorical |
| Bipolar-Mania | M.I.N.I and Young Mania Rating Scale | Yes or No | Categorical |
| Psychosis | M.I.N.I and PANSS | Yes or No | Categorical |
| Apathy | Apathy Evaluation Scale(AES) | Yes or No | Categorical |
| Neurocognitive disorders | Montreal Cognitive Assessment (MoCA) | Yes or No | Categorical |

**Aim 2 variables;** addresses the predictors of neuropsychiatric manifestation in TBI patients at one month which include age, sex, occupation, education level, substance use, intoxication with alcohol during injury, severity of TBI, nature of injury, cause of trauma, area affected, type of lesion, duration of LOC, duration of PTA, history of seizures (See Table 2 in the appendix section for description of variables for aim 2)

**Table 2.** Aim 2.

| Table 2: Description of factors associated with NSP manifestation at one month post TBI |  |  |  |
| --- | --- | --- | --- |
| Variables | Methods of Measurement | Operational definition | Level of Measurements |
| Age | Patient's file report | Age( years) | Continuous |
| Sex | Patient's file report | Male or female | categorical |
| Occupation | Patient's file report | Driver, footballer and others | categorical |
| History of substance use | Patient's file report | Yes or no | categorical |
| History of being intoxicated with alcohol | Patient's file report | Yes or no | categorical |
| **Clinical characteristics** |  |  |  |
| Aphasia | Clinical assessment | Yes or no | categorical |
| Dysarthria | Clinical assessment | Yes or no | categorical |
| Confusion | Clinical assessment | Yes or no | categorical |
| Limb weakness/laterality | Clinical assessment | Yes or no | categorical |
| Seizures/ convulsions | Clinical assessment | Yes or no | categorical |
| **Severity of TBI** |  |  |  |
| Glasgow coma scale | Patient's file report | Mild, moderate, severe | Categorical |
| Duration of LOC | Patient's report | <30minutes,<br>30minutes to 24hrs<br>>24hrs | categorical |
| Duration of PTA | Patient's report | <24hrs<br>1day to 7days<br>>7 days | categorical |
| **Injury characteristics** |  |  |  |
| Nature of injury | Clinical assessment | Open, closed<br>Sharp, blunt<br>Penetrative, non-penetrative | categorical |
| Cause of trauma | Patient's file report | RTA, falls, hit by object | categorical |
| Area affected | CT/MRI | Frontal, temporal, parietal, occipital | categorical |
| Type of lesion | CT/MRI | Hematoma, diffuse axonal injury, cerebral edema | categorical |

**Aim 3 variables;** The variables addresses the cumulative incidence of neuropsychiatric disorders at three and six months following TBI, that is all neuropsychiatric manifestations identified between one and six months, including Depression, Generalized Anxiety Disorder, Bipolar, PTSD, Neurocognitive disorder, Psychosis and Apathy (See table 3 in the appendix section for a description of variables for aim 3)

**Table 3.**
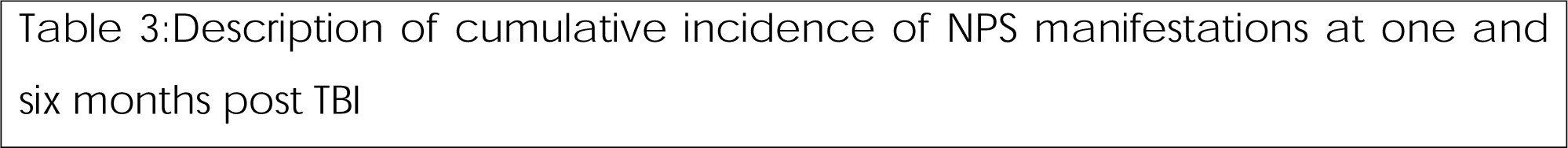
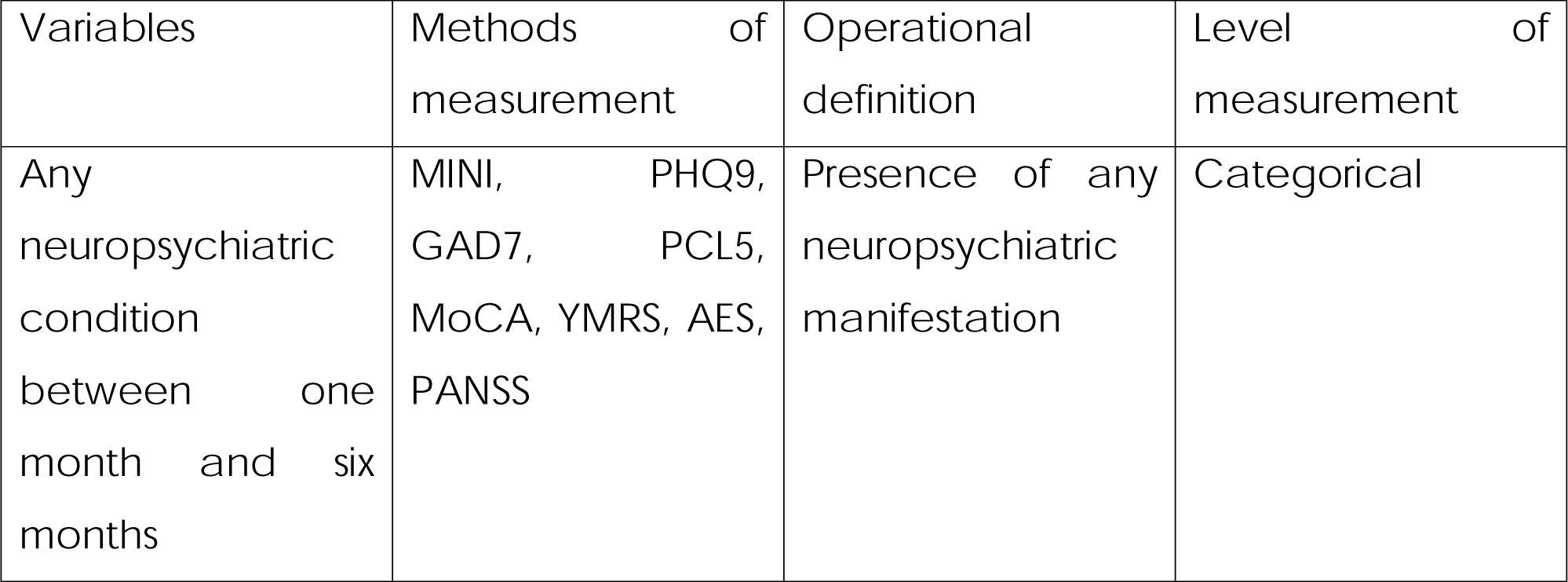
Aim 3.

Description of cumulative incidence of NPS manifestations at one and six months post TBI
| Variables | Methods of measurement | Operational definition | Level of measurement |
| --- | --- | --- | --- |
| Any neuropsychiatric condition between one month and six months | MINI, PHQ9, GAD7, PCL5, MoCA, YMRS, AES, PANSS | Presence of any neuropsychiatric manifestation | Categorical |

**Aim 4 variables**; The variables addresses factors associated with any neuropsychiatric disorder at three and six months which include age, sex, occupation, education level, substance use, intoxication with alcohol during injury, severity of TBI, nature of injury, cause of trauma, area affected, type of lesion, duration of LOC, duration of PTA, history of seizures, presence of any neuropsychiatric manifestation at one and three months, presence of seizures any time during follow up, medications given during follow up period (See Table 4 in the appendix section for description of variables for aim 4)

**Table 4.**
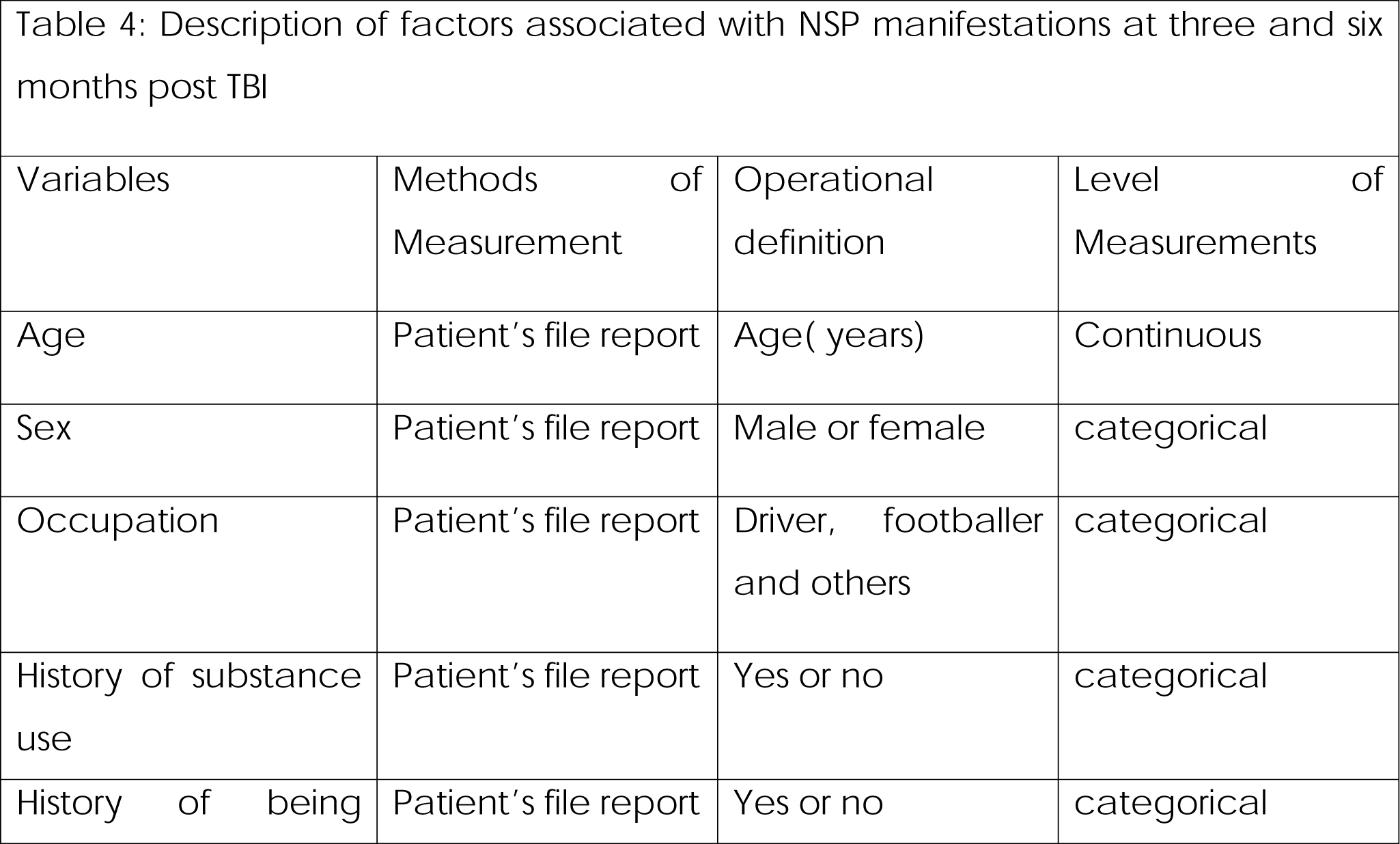
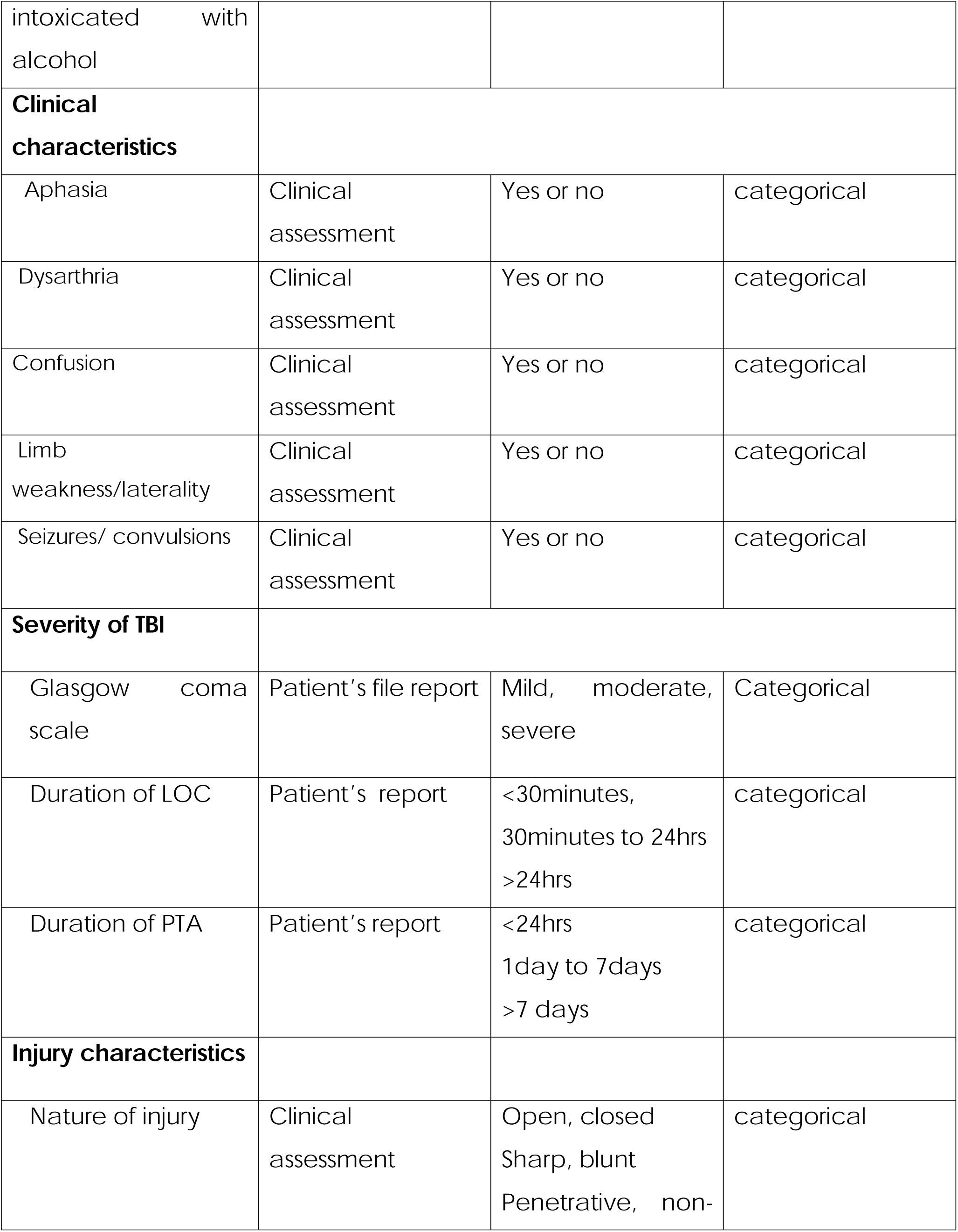
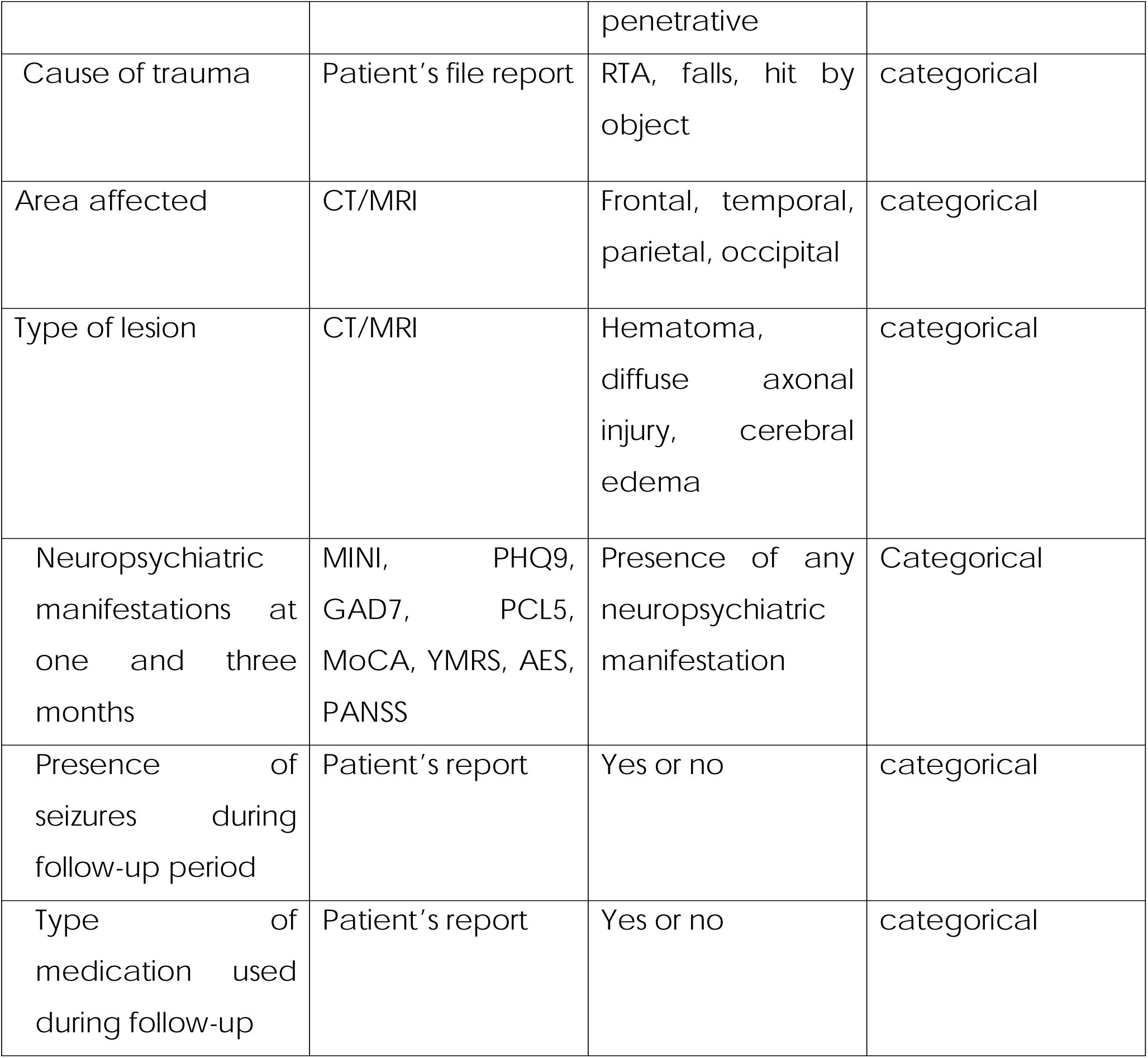
Aim 4.

### Data collection instruments

**Mini International Neuropsychiatric Interview (M.I.N.I.)** will be used to diagnose psychiatric manifestations; It is a diagnostic structured interview which uses the DSM-5 and ICD-10 criteria(Sheehan et al., 1998).We will administer module A(depressive disorder), module C (bipolar disorder),module H (PTSD), module N (Generalized anxiety disorder), and module K (psychosis). It is a useful tool for health workers in diagnosing psychiatric disorders and had accuracy of 91.8% when compared to ICD 10(Chellamuthu, Elangovan, & Karthikeyan, 2017). M.I.N.I is also useful in identifying comorbidities as shown in a study where a third of patients had ≥ 2 psychiatric diagnosis(Pettersson, Modin, Wahlström, Af Winklerfelt Hammarberg, & Krakau, 2018). M.I.N.I which has acceptably high validation and reliability scores and can also be administered much shorter time (mean 18.7 + 11.6 min, median 15 min), after a brief training session, clinicians can use it, and lay interviewers require more extensive training(Amorim, 2000).

**Patient Health Questioner (PHQ-9)** will be used to assess depressive symptoms with scores ranging from 0 to 27, since each of the 9 items (depressed mood, loss of interest in pleasurable activities, weight loss or weight gain, insomnia or hypersomnia, psychomotor agitation or retardation, fatigue or loss of energy, feeling of worthless or guilt, poor concentration and suicidal ideation) matching the Diagnostic and Statistical Manual of Mental Disorders, Fifth Edition (DSM-V) criteria of major depressive disorder. Each item can be scored from 0 (not at all) to 3 (nearly every day)(Manea, Gilbody, & McMillan, 2015). It also measures depression severity with (5-9) categorized as mild, (10-14) as moderate,(15-19) moderately severe, (20-27) severe(Kroenke, Spitzer, & Williams, 2001a). The likely hood of having major depressive disorder is marked at PHQ score ≥ 15(Kroenke, Spitzer, & Williams, 2001b) with sensitivity and specificity of 75% and 76% respectively(Cassin et al., 2013)

**The GAD-7**, a seven-item scale for generalized anxiety disorder, will be used to evaluate anxiety symptoms(feeling anxious, not being able to stop worrying, worrying too much about many things, trouble relaxing, being restless, being easily annoyed or irritable, feeling afraid as if something bad will happen) (Spitzer RL, Kroenke K, Williams JW, & Löwe B, 2006). Response options are “not at all,” “several days,” “more than half the days,” and “nearly every day,” scores are 0, 1, 2, and 3, respectively. In clinical practice and research, GAD-7 is a reliable and accurate tool for screening GAD and measuring the severity of the disorder. A cut-off point of 5 for mild, 10 for moderate and 15 for severe anxiety have demonstrated maximum sensitivity of 89% and specificity of 82%(Spitzer RL et al., 2006).

**The Young Mania Rating Scale (YMRS)** will be used to assess manic symptoms (elevated mood, increased energy or motor activity, increased sexual interest, decrease need for sleep, being easily irritable over talkative, flight of ideas and grandiosity). It is an 11-item interviewer rated scale (Young *et al*., 1978). The four items that are assessed on a 0–8 range are irritability, speech, thought content, and disruptive or violent conduct. The remaining seven categories are graded on a 0–4 scale, with cut-off values of minimal (13), mild (20), moderate (26), and severe (38) manic symptoms. It is a valuable instrument in screening patients with bipolar disorder in the manic phase with the cut-off point of 12.5, sensitivity of 93% and specificity of 96%(Mohammadi et al., 2018).

**Apathy Evaluation Scale (AES)** will be used to assess apathy. It was created by Marin(1991) as a tool for assessing apathy brought on by pathologies of the brain(Kasai, Meguro, & Nakamura, 2014). AES is a reliable and accurate indicator of apathy following a TBI(A. Lane-Brown & Tate, 2009). The scale has 18 items, each evaluated on a Likert scale with four possible responses, and it is measured on a scale from 18 to 72, with higher scores indicating greater apathy (Lee, Gleason, & Umucu, 2020). The optimal trade-off between sensitivity(83%) and specificity(67%) is provided by a score of 36(A. T. Lane-Brown & Tate, 2009).

**Positive and Negative Syndrome Scale (PANSS)**. It is a scale used in medicine to rate the severity of symptoms in people with psychosis(Kay, Fiszbein, & Opler, 1987). Seven Positive Scales, P(delusion, disorganized behavior, hallucination, excitement, persecution/suspiciousness and hostility), seven Negative Scales, N(blunted affect, emotional withdrawal, poor rapport, passive, difficult in abstract thinking, lack of spontaneity, and stereotyped thinking), the remaining sixteen are General Psychopathology Scales, G(somatic complain, anxiety, guilty feeling, tension, mannerism/ posturing, depression, motor retardation, uncooperativeness, unusual thought content, disorientation, poor attention, lack of judgment and insight, disturbance and volition, poor impulse control, preoccupation and active social avoidance) make up the remaining thirty items on the PANSS. The optimum cutoff point of 38.5 has sensitivity of 96% and specificity of 95.9% (Yehya et al., 2016)

**PTSD checklist for DSM- 5 (PLC-5)** will assess PTSD symptoms. Similar to the PCL(Weathers, F. W., Litz, B., Herman, D., Juska, J., & Keane, 1993), the PCL-5 item scores are added up to produce a continuous measure of the severity of PTSD symptoms using DSM 5 criteria for both individual symptom clusters and the entire condition(Jakupcak, 2007). Twenty items make up the PCL-5, each of which is scored on a five-point Likert-type scale and the scores range from "Not at all" (zero) to "Extremely" (four), yielding a symptom severity score that ranges from 0 to 80(Ashbaugh, Houle-Johnson, Herbert, El-Hage, & Brunet, 2016). The cut-off score of 23 has achieved the optimal balance of sensitivity 82% and specificity of 70%(Ibrahim, Ertl, Catani, Ismail, & Neuner, 2018)

**Montreal Cognitive Assessment** will be used to evaluate cognitive impairment (MoCA). It is a rapid cognitive screening instrument that has high sensitivity and specificity for identifying mild cognitive impairment(Coen et al., 2016). MoCA assess visuospatial/executive function which have 5 points, naming have 3 points, memory- 5 points, attention-2 points, language-3 points, abstraction-2 points, and orientation 6 points. A MoCA cutoff score of 23 will be used in this study rather than an initial cutoff score of 26, as it lowers the false positive rate and displays generally improved diagnostic accuracy particularly in less educated population(Carson, Leach, & Murphy, 2018).This meta-analysis have shown that although sensitivity was lower at 23 (83%) than at 26 (94%), but specificity was higher (88% vs. 66%)

### Data analysis plan

All data will be analyzed using the Statistical Package for social Sciences (SPSS) version 25. Descriptive statistics such as mean, median, frequency and standard deviation will describe the baseline characteristics of participants, a logistic regression model will be run to determine the association between predictors and NPS outcome at 1, 3 and 6 months. An Odd ratio > 1 will indicate that there is high likely hood that an independent variable is associated with NPS manifestations or no NPS. Likewise, an odds ratio < 1 will indicate that there is less likely hood that an independent variable is associated with NPS or no NPS manifestation.

Those with p-value less than 0.2 in univariate logistic regression analysis will be computed under multivariable analysis with p-value set at <0.05 level of significance.

## ETHICS AND DISSEMINATION

The ethical clearance has been secured from the institutional Research Review committee and ethical committee of the University of Dodoma with the reference number MA.84/261/12. Permission to conduct the study within the premises of Benjamin Mkapa Hospital (AB.150/293/O1/360) and Dodoma Regional Referral Hospital (PB.22/1307/02/112) were provided by the respective authorities.

The study participants or their relatives will be verbally informed of their participation in the study, and those with the capacity to provide informed consent for participation in the study will be recruited otherwise a close family member or custodian will provide the consent on behalf. The consent form will state clearly the benefit of participation in the study including receiving medical and psychological treatments when needed. The informed consent document will be translated into Swahili, the spoken language of many participants. If the subjects cannot read, the document will be read to them. All consenting subjects will sign the consent forms; if the individual is unable to write, fingerprints will be used instead. The participants will be reassured that they are free to opt out of the study at any point during the study without affecting their access to the routine care and services at the hospitals. Confidentiality will be assured and maintained. Patients diagnosed with any psychiatric illness will be referred for further assessment and treatment. Patients or the public WERE NOT involved in the design, or conduct, or reporting, or dissemination plans of our research

## Data Availability

All data produced in the present study are available upon reasonable request to the authors

## Author Contributions

Conceptualization: S A, A N.

Data curation: S A.

Formal analysis: S A, A N.

Investigation: S A, A N.

Methodology: S A, A N.

Supervision: S A, A N.

Writing – original draft: S A.

Writing – review & editing: S A, A N.

## Funding statement

The data collection of this project is partially funded by The Ministry of Health Zanzibar.

## Data availability statement

Data will be available and shared as per agreement of terms and conditions once the data collection is completed

## Competing interest statement

The authors declare there is no conflict of interest.

## Notes

### Competing Interest Statement

The authors have declared no competing interest.

### Funding Statement

No funding was acquired for this manuscript.

### Author Declarations

The Institutional Research Review Ethics Committee of the University of Dodoma.

